# HIV and risk of COVID-19 death: a population cohort study from the Western Cape Province, South Africa

**DOI:** 10.1101/2020.07.02.20145185

**Authors:** Mary-Ann Davies

## Abstract

**Background:** The effect of HIV co-infection on COVID-19 outcomes in sub-Saharan Africa is unknown.

**Methods:** We conducted a population cohort study using linked data from adults attending public sector health facilities in the Western Cape, South Africa. We used Cox-proportional hazards models adjusted for age, sex, location and comorbidities to examine the association between HIV and COVID-19 death among (i) public sector “active patients” (≥1 health visit in the 3 years before March 2020), (ii) laboratory-diagnosed COVID-19 cases and (iii) hospitalized COVID-19 cases. COVID-19 was diagnosed with SARS-CoV-2 PCR tests. We calculated the standardized mortality ratio (SMR) for COVID-19 comparing HIV positive vs. negative adults using modelled population estimates.

**Results:** Among 3,460,932 public sector patients (16% HIV positive), 22,308 were diagnosed with COVID-19, of whom 625 died. In adjusted analysis, HIV increased risk of COVID-19 mortality (adjusted hazard ratio [aHR]:2.14; 95% confidence interval [CI]:1.70; 2.70), with similar risks across strata of viral load and immunosuppression. increased HIV-associated risk of COVID-19 death remained when restricting to COVID-19 cases (aHR:1.70; 95%CI:132; 2.18) or hospitalized cases (aHR:1.45; 95%CI:1.14; 1.84). Current and previous tuberculosis also increased COVID-19 mortality risk (aHR [95%CI]:2.70 [1.81; 4.04] and 1.51 [1.18; 1.93] respectively). The SMR for COVID-19 death associated with HIV was 2.39 (95% CI:1.96; 2.86); population attributable fraction 8.5% (95%CI:6.1; 11.1).

**Conclusion:** HIV was associated with a doubling of COVID-19 mortality risk. While our findings may over-estimate the HIV-associated risk COVID-19 death due to residual confounding, people with HIV should be considered a high-risk group for COVID-19 management.

## Introduction

The effects of the intersecting pandemics of HIV, tuberculosis and coronavirus disease-19 (COVID-19) in sub-Saharan Africa are unknown. Studies to date suggest no increased risk of adverse outcomes in HIV co-infected patients, but these are small case series from Europe and North America, often limited to hospitalized patients, and may not be relevant to sub-Saharan Africa where people living with HIV (PLWH) are younger with different comorbidities, frequently including tuberculosis.^1-7^ PLWH may experience more severe COVID-19 disease due to HIV-related immune suppression, which may be exacerbated by transient immune deficiency from coronaviruses.^8,9^ In support of this hypothesis a large UK cohort study reported increased risk of COVID-19 death with immunosuppressive comorbidity, including PLWH.^10^ Two factors may reduce risk of severe COVID-19 in PLWH: dysfunctional cellular immunity could protect against the cytokine storm,^11,12^ and some antiretroviral drugs (tenofovir and lopinavir-ritonavir) have *in vitro* activity against coronaviruses, with better outcomes reported for PLWH receiving tenofovir disoproxil fumarate (TDF) vs. other antiretrovirals.^12,13^

It is important to establish if HIV and tuberculosis increase risk of COVID-19 death so that patients with these conditions can be provided with augmented prevention and potential therapeutic interventions. We used linked data from adults attending public sector health facilities in the Western Cape Province, South Africa, to identify factors associated with COVID-19 death, including HIV and tuberculosis.

## Methods

### Study design

We conducted a cohort study using de-identified data from the Western Cape Provincial Health Data Centre (WCPHDC) of public sector patients aged ≥20 years with documented sex and not known to have died before March 1, 2020 (before the first diagnosed COVID-19 case in South Africa and several weeks before the first COVID-19 death) and included all follow up through June 9, 2020. The outcome was COVID-19 death. Our main analysis examined risk of COVID-19 death in the general population, so all patients were included regardless of SARS-CoV-2 testing. The study was approved by the University of Cape Town and Stellenbosch University Health Research Ethics Committees and the Western Cape Province Department of Health. Individual informed consent requirement was waived for this secondary analysis of de-identified data.

### Study population and data sources

The Western Cape has nearly 7 million inhabitants, of whom ∼520,000 are PLWH with more than 90% of them dependent on public sector health services. The WCPHDC has been described in detail.^14^ Briefly, WCPHDC consolidates administrative, laboratory, and pharmacy data from routine electronic clinical information systems used in all public sector health facilities with linkage through a unique identifier. Multiple data sources are triangulated to enumerate health conditions such as diabetes mellitus (“diabetes”), hypertension, tuberculosis and HIV, with high or moderate certainty evidence assigned for each inferred condition (Supplementary Table 1). Patients are deemed to be PLWH (high certainty) if they have a positive HIV diagnostic test and/or HIV-RNA test and/or have received triple antiretroviral therapy (ART) and/or are registered in the HIV chronic disease management system; moderate certainty is assigned for those with only a CD4 count measure and/or two antiretroviral drugs prescribed and/or ICD-10 diagnosis code of HIV. HIV testing coverage is high; >90% of PLWH in the province know their HIV diagnosis.^15^ Comorbidities were based on high or moderate certainty evidence for the main analysis and restricted to high certainty evidence in sensitivity analyses. The virologic, immunologic and ART status of PLWH on March 1, 2020 was categorized, based on most recent measures, as “confirmed virologically suppressed on ART” (HIV-RNA<1000 copies/ml in last 15 months and ART dispensed in last 6 months), “likely virologically suppressed on ART” (HIV-RNA <1000 copies/ml 15-24 months previously or HIV-RNA <1000 copies/ml >24 months previously if ART dispensed in last 6 months), “viraemic or immunosuppressed” (HIV-RNA >1000 copies/ml in last 15 months or CD4 count <200 cells/µl within 18 months before March 2020) or “unknown”. Among hospitalized COVID-19 cases we examined the association between COVID-19 death and CD4 count at COVID-19 diagnosis/admission. Until January 2020, adult first-line ART was TDF plus emtricitabine/lamivudine plus efavirenz, with abacavir replacing TDF for patients with kidney disease; zidovudine plus emtricitabine/lamivudine plus a protease inhibitor was used for second-line ART. Dolutegravir was introduced in first and second-line since January 2020. Diabetic control was categorized according to glycosylated haemoglobin (HbA1c) measurement within the last 2 years as <7% (controlled); 7-8.9% (poorly controlled), ≥9% (uncontrolled).

### COVID-19 diagnosis

All COVID-19 diagnoses were based on positive SARS-CoV-2 PCR tests. Testing was available for all patients with COVID-19 symptoms until June 1, 2020; thereafter public sector laboratory testing was restricted to patients requiring admission or older than 55 years or with comorbidities, due to a temporary limit in testing capacity. Asymptomatic contacts of cases were not routinely tested. Hospital admissions and all deaths in SARS-CoV-2 positive cases are recorded and reviewed daily in the WCPHDC.

### Statistical analysis

We used Cox-proportional hazards models adjusted for age, sex and other comorbidities to examine the association between HIV and COVID-19 death among (i) all public sector patients with ≥1 health visit in the 3 years before March 1, 2020 (considered “active patients”), (ii) laboratory-diagnosed COVID-19 cases and (iii) hospitalized COVID-19 cases. We adjusted for location within Cape Town vs. rest of the province and subdistrict of residence for Cape Town patients to account for geographical variation in infection rates and as a proxy for socio-economic status. Patients were censored on the date of death if deceased without a COVID-19 diagnosis, or on June 9, 2020, whichever was earliest. Database closure was 7 days later to allow death reporting delays. For the analysis of COVID-19 death in laboratory-diagnosed cases we included cases diagnosed before June 1, 2020 when testing was available for all patients with COVID-19 symptoms, but included all patients diagnosed by June 9, 2020 in a sensitivity analysis. The proportional-hazard assumption was assessed with Schoenfeld residuals. All analyses were conducted using Stata 15.1.

We also calculated the standardized mortality ratio (SMR) of the actual number of COVID-19 deaths in people with HIV vs. the number that would be expected if PLWH had the same risk of COVID-19 death as HIV-negative people of the same age and sex. We used data on the age, sex and HIV status of all COVID-19 deaths (public and private sector) and the Thembisa Western Cape HIV model to estimate the Western Cape population size and HIV prevalence, by age and sex, in 2020.^16^ We used 1000 bootstrap replications to calculate 95% confidence intervals (CI) for the SMR (See Supplementary Appendix 2).

Since individual socio-economic status and some comorbidities are not recorded in WCPHDC, including body mass index (BMI), we calculated E-values to determine the minimum strength of association that an unmeasured confounder (such as raised BMI or differences in socio-economic status) would need to have with both HIV positive status and COVID-19 death to fully account for any association between HIV and COVID-19 death.^17^ We conducted quantitative bias analysis to assess the impact of potential confounding by obesity.

## Results

### Patient characteristics

Among 3,460,932 “active patients” aged ≥20 years or older on March 1, 2020, 22,308 were diagnosed with COVID-19, of whom 625 (2.8%) died (Table 1). The proportion of men was lower among COVID-19 cases vs. non-cases (31% vs. 42%), likely due to initial cases being among essential workers in sectors employing predominantly women. Diabetes and hypertension were common in all patients, with higher prevalence among COVID-19 cases than non-cases, (diabetes:14% vs. 8%; hypertension: 23% vs 16%), while proportions of other comorbidities were similar. COVID-19 deceased cases were older than surviving cases (median age [interquartile range] 63 years [54-71] vs. 37 [30-48]); several comorbidities were more common among deceased patients.

**Table 1:** Characteristics of (i) Western Cape “active patients” aged ≥20 years in public sector (public sector health care visit in last 3 years before March 1, 2020) according to COVID-19 outcome (ii) COVID-19 cases in “active patients” and (iii) hospitalized COVID-19 cases in “active patients”.

|  | All public sector patients |  |  | All public-sector SARS-CoV-2 cases diagnosed before 1 June 2020* |  | Hospitalized public-sector SARS-CoV-2 cases |  |
| --- | --- | --- | --- | --- | --- | --- | --- |
|  | No diagnosed COVID-19<br>n=3,438,624 | COVID-19 not deceased<br>n=21,683 | COVID-19 deceased<br>n=625 | COVID-19 not deceased<br>n=14,693 | COVID-19 deceased<br>n=510 | COVID-19 not deceased<br>n=2,428 | COVID-19 deceased<br>n=550 |
| **Sex** |  |  |  |  |  |  |  |
| female | 1,983,480 (58%) | 14,916 (69%) | 340 (54%) | 10,388 (71%) | 281 (55%) | 1,546 (64%) | 302 (55%) |
| male | 1,455,144 (42%) | 6,767 (31%) | 285 (46%) | 4,305 (29%) | 229 (45%) | 882 (36%) | 248 (49%) |
| **Age** |  |  |  |  |  |  |  |
| 20-39 years | 1,913,786 (56%) | 11,64 (54%) | 46 (7%) | 8,653 (59%) | 35 (7%) | 804 (33%) | 45 (8%) |
| 40-49 years | 604,976 (18%) | 4,515 (21%) | 63 (10%) | 2,991 (20%) | 51 (10%) | 472 (19%) | 56 (10%) |
| 50-59 years | 447,739 (13%) | 3,227 (15%) | 162 (26%) | 1,881 (13%) | 137 (27%) | 523 (21%) | 142 (26%) |
| 60-69 years | 276,082 (8%) | 1,423 (7%) | 178 (28%) | 739 (5%) | 150 (29%) | 360 (15%) | 158 (29%) |
| $\geq 70$ years | 196,035 (6%) | 878 (4%) | 176 (28%) | 429 (3%) | 137 (27%) | 269 (11%) | 149 (27%) |
| **Diabetes** |  |  |  |  |  |  |  |
| none | 3,177,088 (92%) | 18,581 (86%) | 253 (40%) | 12,921 (88%) | 212 (42%) | 1,549 (64%) | 225 (41%) |
| diabetes HbA1c <7% | 45,054 (1%) | 491 (2%) | 58 (9%) | 278 (2%) | 43 (8%) | 149 (6%) | 54 (10%) |
| diabetes HbA1c 7 - 8.9% | 47,211 (1%) | 582 (3%) | 94 (15%) | 309 (2%) | 79 (15%) | 164 (7%) | 85 (15%) |
| diabetes HbA1c $\geq 9\%$ | 65,639 (2%) | 1,086 (5%) | 158 (25%) | 640 (4%) | 125 (25%) | 375 (15%) | 136 (25%) |
| diabetes no HbA1c measurement | 103,632 (3%) | 943 (4%) | 62 (10%) | 545 (4%) | 51 (10%) | 191 (8%) | 50 (9%) |
| **Other non-communicable diseases** |  |  |  |  |  |  |  |
| hypertension | 563,908 (16%) | 4,910 (23%) | 362 (58%) | 2,972 (20%) | 293 (57%) | 948 (39%) | 319 (58%) |
| chronic kidney disease | 61,667 (2%) | 494 (2%) | 111 (18%) | 250 (2%) | 92 (18%) | 168 (7%) | 101 (18%) |
| chronic pulmonary disease / asthma | 192,587 (6%) | 1,577 (7%) | 84 (13%) | 949 (6%) | 77 (15%) | 355 (15%) | 78 (14%) |
| **Tuberculosis** |  |  |  |  |  |  |  |
| never tuberculosis | 3,097,483 (90%) | 19,668 (91%) | 512 (82%) | 13,330 (91%) | 414 (81%) | 2061 (85%) | 448 (81%) |
| previous tuberculosis | 286,889 (8%) | 1,698 (8%) | 87 (14%) | 1,180 (8%) | 74 (15%) | 244 (10%) | 77 (14%) |
| current tuberculosis | 54252 (2%) | 317 (1%) | 26 (4%) | 213 (1%) | 22(4%) | 123 (5%) | 25 (5%) |
| **HIV** |  |  |  |  |  |  |  |
| negative | 2,902,050 (84%) | 17,820 (82%) | 510 (82%) | 11,893 (81%) | 415 (81%) | 1,932 (80%) | 445 (81%) |
| positive | 536,574 (16%) | 3,863 (18%) | 115 (18%) | 2,800 (19%) | 95 (19%) | 496 (20%) | 105 (19%) |
| **VL <1000 copies/ml (last 15 mo) & ART script (last 6 mo)** | 240,048 (45%) | 2,332 (60%) | 71 (62%) | 1,726 (62%) | 57 (60%) | 289 (58%) | 68 (65%) |
| **VL <1000 copies/ml (2yr to 15 mo prior)**<br>**OR ART script (last 6 mo) & VL <1000 copies/ml > 2yr prior** | 68,871 (13%) | 409 (11%) | 11 (10%) | 283 (10%) | 9 (9%) | 46 (9%) | 11 (10%) |
| **VL $\geq 1000$ copies/ml (last 15 mo) or CD4 <200 cells/μl (last 18 mo)** | 40,974 (8%) | 209 (5%) | 12 (10%) | 150 (5%) | 10 (11%) | 58 (12%) | 9 (9%) |
| **No VL (last 15 mo); CD4 $\geq 200$ cells/μl or unknown (last 18 mo)** | 186,681 (35%) | 913 (24%) | 21 (18%) | 641 (23%) | 19 (20%) | 103 (21%) | 17 (16%) |

\*Cases analysis limited to cases diagnosed before 1 June 2020 when testing criteria changed with public sector tests being limited to patients >55 year of age or with comorbidities; Note: Column percentages may add up to >100% due to rounding; HbA1c glycosylated haemoglobin; VL viral load; mo months; yr years; ART antiretroviral therapy

### Patients with and without HIV

Although the proportion of PLWH was similar among surviving and deceased COVID-19 cases, a greater proportion of COVID-19 deaths were in patients aged <50 years in those with vs. without HIV (39% vs 13%) (Table 2). A substantial proportion of COVID-19 deceased PLWH had diabetes (50%) and hypertension (42%), however these conditions were more common in deceased people without HIV. Current and previous tuberculosis were more frequent in PLWH irrespective of COVID-19; a high proportion of COVID-19 deceased cases with HIV had current (14%) and/or previous tuberculosis (37%).

**Table 2:** Characteristics of (i) Western Cape “active patients” ≥20 years of age in public sector (public sector health care visit in last 3 years) with and without HIV according to COVID-19 outcome.

|  | Public sector patients with HIV |  |  | Public sector patients without HIV |  |  |
| --- | --- | --- | --- | --- | --- | --- |
|  | No diagnosed<br>COVID-19<br>n=536,574 | COVID-19 not<br>deceased<br>n=3,863 | COVID-19<br>deceased<br>n=115 | No diagnosed<br>COVID-19<br>n=2,902,050 | COVID-19 not<br>deceased<br>n=17,820 | COVID-19<br>deceased<br>n=510 |
| **Sex** |  |  |  |  |  |  |
| female | 356,356 (66%) | 3,039 (79%) | 62 (54%) | 1,627,124 (56%) | 11,877 (67%) | 278 (55%) |
| male | 180,218 (34%) | 824 (21%) | 53 (46%) | 1,274,926 (44%) | 5,943 (34%) | 232 (45%) |
| **Age** |  |  |  |  |  |  |
| 20-39 years | 310,551 (58%) | 2,187 (57%) | 17 (15%) | 1,603,235 (55%) | 9,453 (53%) | 29 (6%) |
| 40-49 years | 147,344 (27%) | 1,136 (29%) | 28 (24%) | 457,632 (16%) | 3,379 (19%) | 35 (7%) |
| 50-59 years | 59,345 (11%) | 418 (11%) | 40 (35%) | 388,394 (13%) | 2,809 (16%) | 122 (24%) |
| 60-69 years | 15,856 (3%) | 98 (3%) | 21 (18%) | 260,226 (9%) | 1,325 (7%) | 157 (31%) |
| $\geq 70$ years | 3,473 (1%) | 24 (1%) | 9 (8%) | 192,562 (7%) | 854 (5%) | 167 (33%) |
| **Diabetes** |  |  |  |  |  |  |
| none | 517,609 (96%) | 3,491 (90%) | 57 (50%) | 2,659,479 (92%) | 15,090 (85%) | 196 (38%) |
| diabetes HbA1c <7% | 3,493 (1%) | 65 (2%) | 8 (7%) | 41,561 (1%) | 426 (2%) | 50 (10%) |
| diabetes HbA1c 7 - 8.9% | 2,998 (1%) | 77 (2%) | 16 (14%) | 44,213 (2%) | 505 (3%) | 78 (15%) |
| diabetes HbA1c $\geq 9\%$ | 4,562 (1%) | 126 (3%) | 25 (22%) | 61,077 (2%) | 960 (5%) | 133 (26%) |
| diabetes no HbA1c measurement | 7,912 (1%) | 104 (3%) | 9 (8%) | 95,720 (3%) | 839 (5%) | 53 (10%) |
| **Other non-communicable diseases** |  |  |  |  |  |  |
| hypertension | 62,676 (12%) | 692 (18%) | 48 (42%) | 501,232 (18%) | 4,218 (24%) | 314 (62%) |
| chronic kidney disease | 6,348 (1%) | 82 (2%) | 21 (18%) | 55,319 (2%) | 412 (2%) | 90 (18%) |
| chronic pulmonary disease / asthma | 23,501 (4%) | 218 (6%) | 10 (9%) | 169,086 (6%) | 1,359 (8%) | 74 (15%) |
| **Tuberculosis** |  |  |  |  |  |  |
| previous tuberculosis | 129,259 (24%) | 864 (22%) | 42 (37%) | 157,630 (5%) | 834 (5%) | 45 (9%) |
| current tuberculosis | 24,357 (5%) | 172 (4%) | 16 (14%) | 29,895 (1%) | 145 (1%) | 10 (2%) |

Note: Column percentages may add up to >100% due to rounding; HbA1c glycosylated haemoglobin

### COVID-19 death in all public sector patients

Among all public sector patients, the probability of COVID-19 death by 100 days since March 1, 2020 was 180/million (95% confidence interval [CI]: 167; 196). COVID-19 death was associated with male sex, increasing age, diabetes (with higher risk with elevated HbA1c), hypertension and chronic kidney disease (Table 3). Current tuberculosis increased the hazard of COVID-19 death (adjusted hazard ratio [aHR]:2.70; 95%CI:1.81; 4.04) with a smaller increased risk for previous tuberculosis (aHR:1.51; 95%CI:1.18; 1.93). The increased risk of COVID-19 death with current tuberculosis was present for both rifampicin-sensitive and resistant disease during intensive phase treatment (Supplementary Table 3).

**Table 3:**
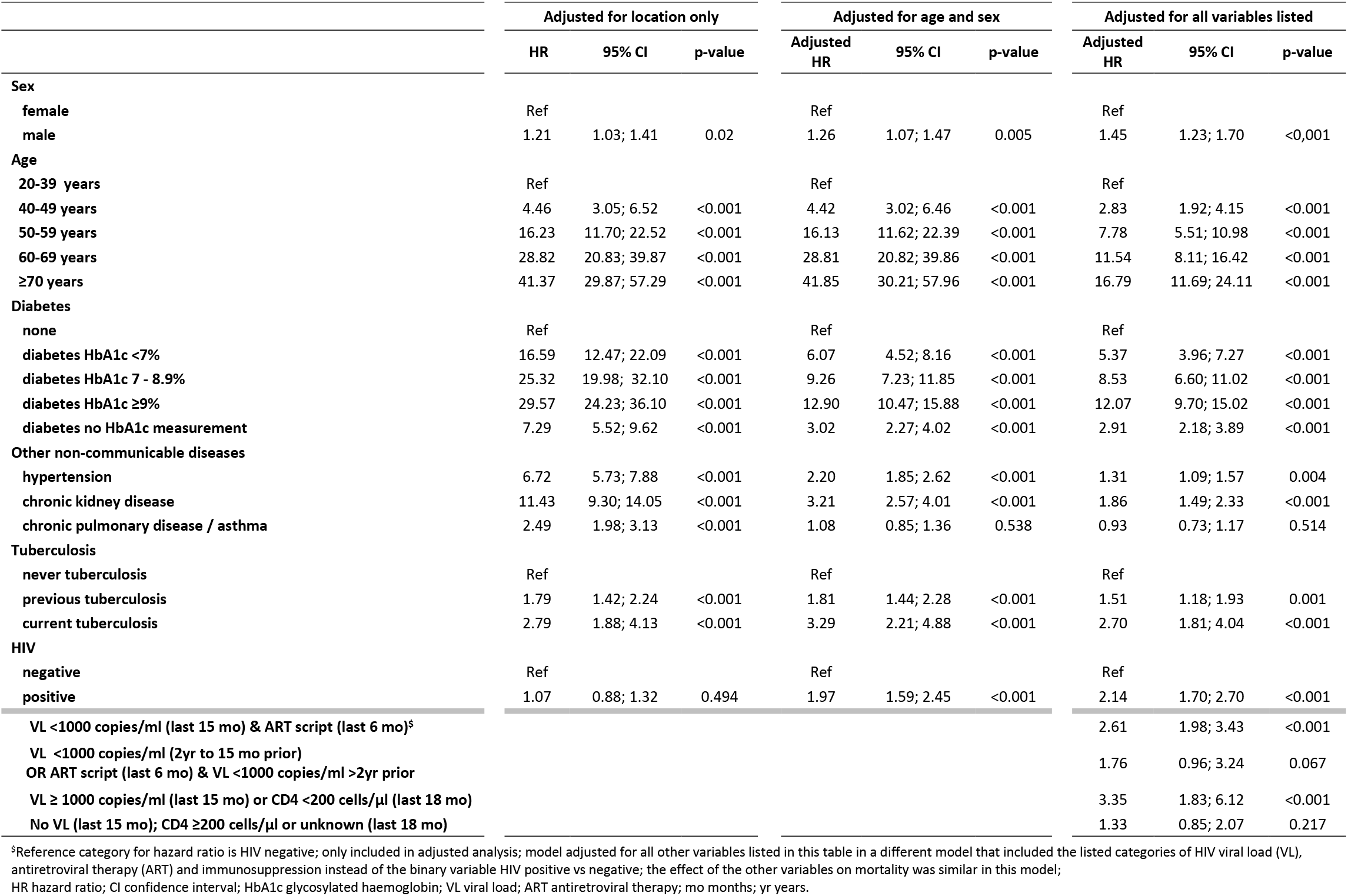
Univariate and multivariate hazard ratios (HRs) and 95% confidence intervals (CI) for associations with COVID-19 death from March 1 to June 9, 2020, among all public sector patients ≥20 years with a public sector health visit in the previous 3 years (n=3,460,932), using Cox-proportional hazards models

The hazard of COVID-19 death adjusted for age and sex was increased in PLWH compared to HIV-negative patients, and the association increased further when adjusting for all recorded comorbidities (aHR:2.14; 95%CI:1.70; 2.70). Associations with most comorbidities increased when restricting to those with high certainty comorbidity evidence (Supplementary Table 4). Increased mortality risk was similar in PLWH irrespective of viraemia/immunosuppression, but there were few viraemic or immunosuppressed patients, as reflected in the wide CIs for the hazard ratio in different groups.

### Death in COVID-19 cases and hospitalized patients

Among the 15,203 COVID-19 cases diagnosed before June 1, 2020, mortality was increased in men and older patients (Table 4; Figure 1). Although the increased risk of death associated with all comorbidities was attenuated compared to the population analysis, PLWH still had an increased hazard of death compared to HIV negative COVID-19 cases (aHR:1.70; 95%CI:1.32; 2.18), with similar results when including patients diagnosed after the change in testing criteria (Supplementary Table 5). Among COVID-19 cases in PLWH on ART, mortality was lower in patients on TDF (vs. abacavir/zidovudine) (aHR: 0.42; 95%CI: 0.22;0.78) with no difference for other antiretrovirals. Among hospitalized COVID-19 cases, the effects of all mortality risk factors including HIV were attenuated, but HIV remained associated with death (aHR:1.45; 95%CI:1.14; 1.84). Among hospitalized patients, CD4 <200 cells/µl at COVID-19 diagnosis or admission was associated with COVID-19 death (aHR vs HIV negative: 2.36; 95%CI:1.47; 3.78).

**Table 4:**
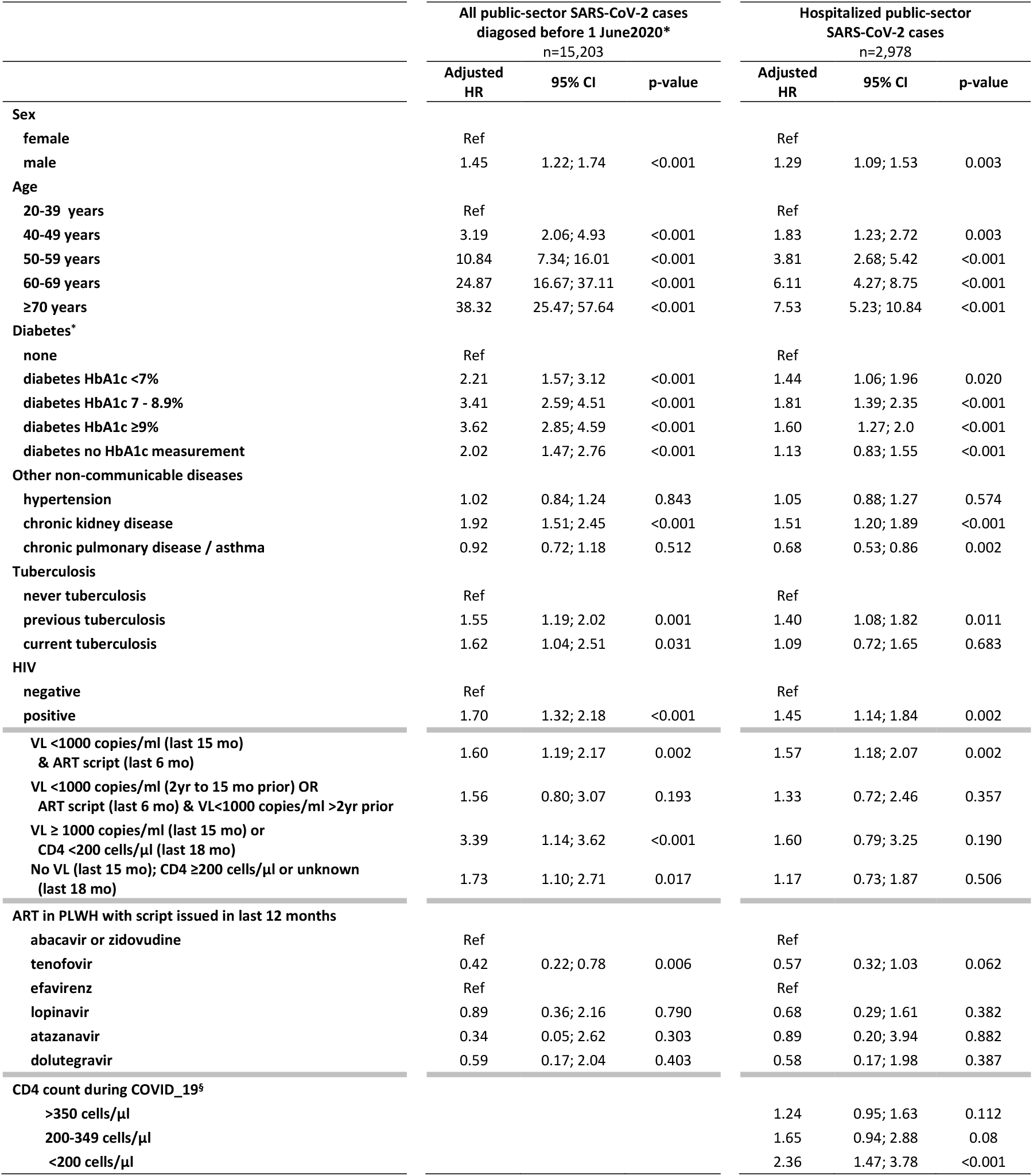
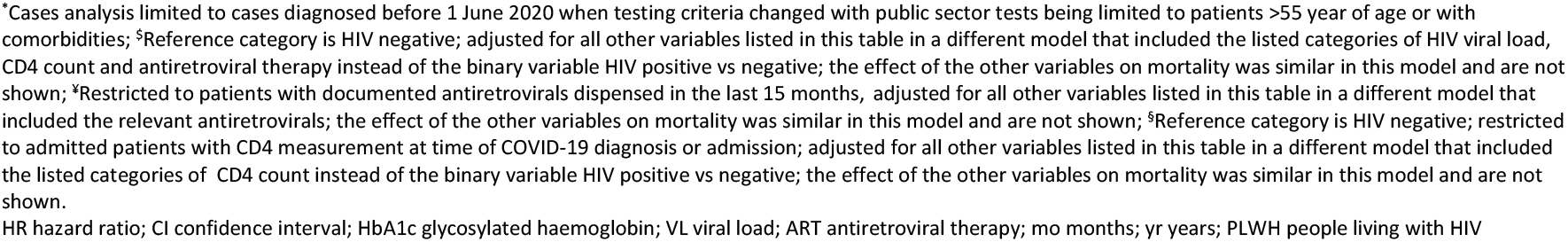
Multivariate hazard ratios (HRs) and 95% confidence intervals (CI) for associations with COVID-19 death from Cox-proportional hazards models among (i) all adult COVID-19 cases diagnosed before June 1, 2020 (n=15,203) and (ii) all hospitalized adult COVID-19 cases (n=2,978).

|  | All public-sector SARS-CoV-2 cases<br>diagnosed before 1 June 2020*<br>n=15,203 |  |  | Hospitalized public-sector<br>SARS-CoV-2 cases<br>n=2,978 |  |  |
| --- | --- | --- | --- | --- | --- | --- |
|  | Adjusted<br>HR | 95% CI | p-value | Adjusted<br>HR | 95% CI | p-value |
| **Sex** |  |  |  |  |  |  |
| female | Ref |  |  | Ref |  |  |
| male | 1.45 | 1.22; 1.74 | <0.001 | 1.29 | 1.09; 1.53 | 0.003 |
| **Age** |  |  |  |  |  |  |
| 20-39 years | Ref |  |  | Ref |  |  |
| 40-49 years | 3.19 | 2.06; 4.93 | <0.001 | 1.83 | 1.23; 2.72 | 0.003 |
| 50-59 years | 10.84 | 7.34; 16.01 | <0.001 | 3.81 | 2.68; 5.42 | <0.001 |
| 60-69 years | 24.87 | 16.67; 37.11 | <0.001 | 6.11 | 4.27; 8.75 | <0.001 |
| ≥70 years | 38.32 | 25.47; 57.64 | <0.001 | 7.53 | 5.23; 10.84 | <0.001 |
| **Diabetes\*** |  |  |  |  |  |  |
| none | Ref |  |  | Ref |  |  |
| diabetes HbA1c <7% | 2.21 | 1.57; 3.12 | <0.001 | 1.44 | 1.06; 1.96 | 0.020 |
| diabetes HbA1c 7 - 8.9% | 3.41 | 2.59; 4.51 | <0.001 | 1.81 | 1.39; 2.35 | <0.001 |
| diabetes HbA1c ≥9% | 3.62 | 2.85; 4.59 | <0.001 | 1.60 | 1.27; 2.0 | <0.001 |
| diabetes no HbA1c measurement | 2.02 | 1.47; 2.76 | <0.001 | 1.13 | 0.83; 1.55 | <0.001 |
| **Other non-communicable diseases** |  |  |  |  |  |  |
| hypertension | 1.02 | 0.84; 1.24 | 0.843 | 1.05 | 0.88; 1.27 | 0.574 |
| chronic kidney disease | 1.92 | 1.51; 2.45 | <0.001 | 1.51 | 1.20; 1.89 | <0.001 |
| chronic pulmonary disease / asthma | 0.92 | 0.72; 1.18 | 0.512 | 0.68 | 0.53; 0.86 | 0.002 |
| **Tuberculosis** |  |  |  |  |  |  |
| never tuberculosis | Ref |  |  | Ref |  |  |
| previous tuberculosis | 1.55 | 1.19; 2.02 | 0.001 | 1.40 | 1.08; 1.82 | 0.011 |
| current tuberculosis | 1.62 | 1.04; 2.51 | 0.031 | 1.09 | 0.72; 1.65 | 0.683 |
| **HIV** |  |  |  |  |  |  |
| negative | Ref |  |  | Ref |  |  |
| positive | 1.70 | 1.32; 2.18 | <0.001 | 1.45 | 1.14; 1.84 | 0.002 |
| **VL <1000 copies/ml (last 15 mo)**<br>**& ART script (last 6 mo)** | 1.60 | 1.19; 2.17 | 0.002 | 1.57 | 1.18; 2.07 | 0.002 |
| **VL <1000 copies/ml (2yr to 15 mo prior) OR**<br>**ART script (last 6 mo) & VL<1000 copies/ml >2yr prior** | 1.56 | 0.80; 3.07 | 0.193 | 1.33 | 0.72; 2.46 | 0.357 |
| **VL ≥ 1000 copies/ml (last 15 mo) or**<br>**CD4 <200 cells/μl (last 18 mo)** | 3.39 | 1.14; 3.62 | <0.001 | 1.60 | 0.79; 3.25 | 0.190 |
| **No VL (last 15 mo); CD4 ≥200 cells/μl or unknown**<br>**(last 18 mo)** | 1.73 | 1.10; 2.71 | 0.017 | 1.17 | 0.73; 1.87 | 0.506 |
| **ART in PLWH with script issued in last 12 months** |  |  |  |  |  |  |
| abacavir or zidovudine | Ref |  |  | Ref |  |  |
| tenofovir | 0.42 | 0.22; 0.78 | 0.006 | 0.57 | 0.32; 1.03 | 0.062 |
| efavirenz | Ref |  |  | Ref |  |  |
| lopinavir | 0.89 | 0.36; 2.16 | 0.790 | 0.68 | 0.29; 1.61 | 0.382 |
| atazanavir | 0.34 | 0.05; 2.62 | 0.303 | 0.89 | 0.20; 3.94 | 0.882 |
| dolutegravir | 0.59 | 0.17; 2.04 | 0.403 | 0.58 | 0.17; 1.98 | 0.387 |
| **CD4 count during COVID_19<sup>§</sup>** |  |  |  |  |  |  |
| >350 cells/μl |  |  |  | 1.24 | 0.95; 1.63 | 0.112 |
| 200-349 cells/μl |  |  |  | 1.65 | 0.94; 2.88 | 0.08 |
| <200 cells/μl |  |  |  | 2.36 | 1.47; 3.78 | <0.001 |

\*Cases analysis limited to cases diagnosed before 1 June 2020 when testing criteria changed with public sector tests being limited to patients >55 year of age or with comorbidities; <sup>5</sup>Reference category is HIV negative; adjusted for all other variables listed in this table in a different model that included the listed categories of HIV viral load, CD4 count and antiretroviral therapy instead of the binary variable HIV positive vs negative; the effect of the other variables on mortality was similar in this model and are not shown; <sup>6</sup>Restricted to patients with documented antiretrovirals dispensed in the last 15 months, adjusted for all other variables listed in this table in a different model that included the relevant antiretrovirals; the effect of the other variables on mortality was similar in this model and are not shown; <sup>8</sup>Reference category is HIV negative; restricted to admitted patients with CD4 measurement at time of COVID-19 diagnosis or admission; adjusted for all other variables listed in this table in a different model that included the listed categories of CD4 count instead of the binary variable HIV positive vs negative; the effect of the other variables on mortality was similar in this model and are not shown.
HR hazard ratio; CI confidence interval; HbA1c glycosylated haemoglobin; VL viral load; ART antiretroviral therapy; mo months; yr years; PLWH people living with HIV

**Figure 1:**
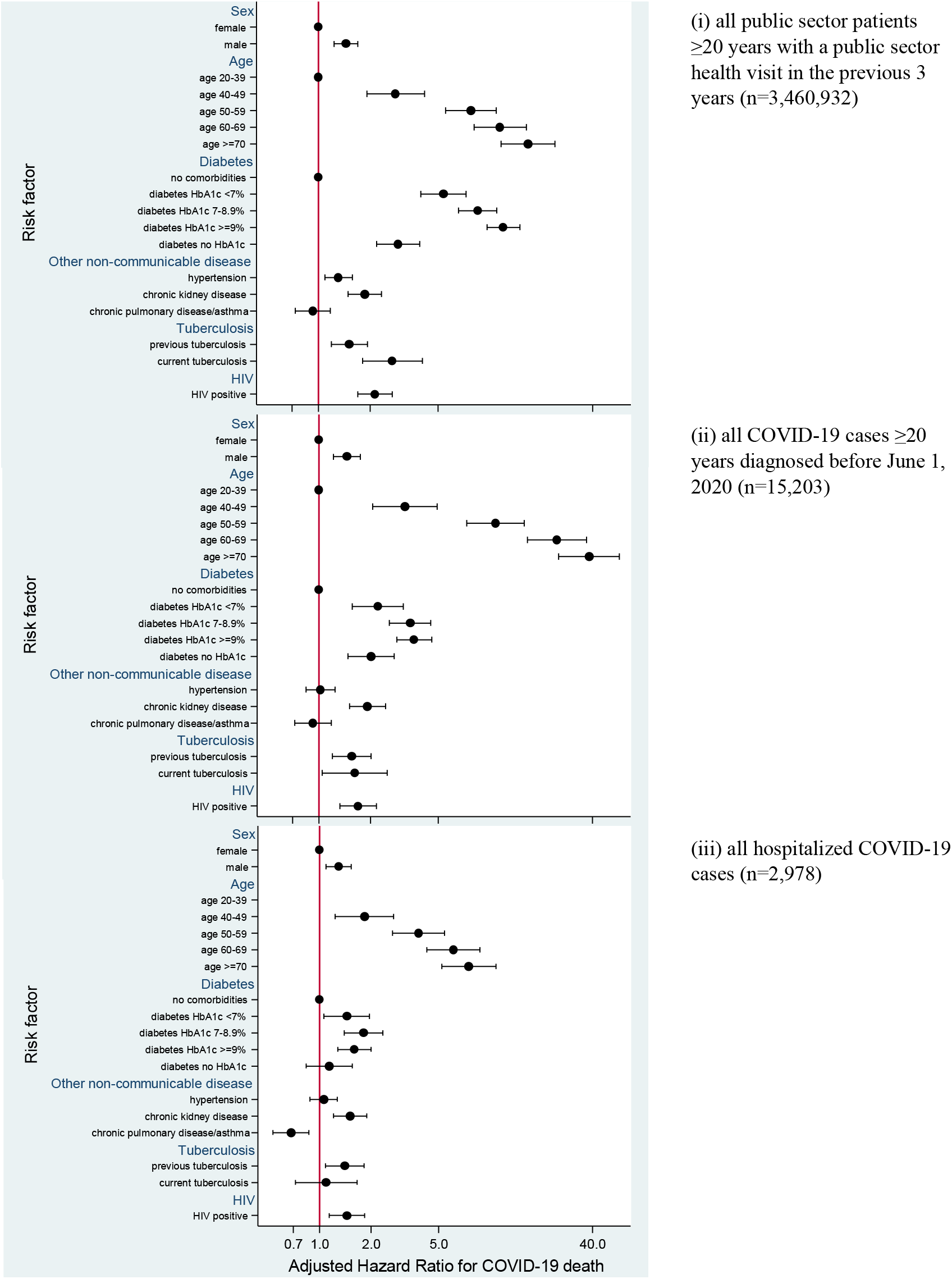
Comparison of adjusted hazard ratios (HR) and 95% confidence intervals (CI) for associations with COVID-19 death from Cox-proportional hazards models among (i) all public sector patients ≥20 years with a public sector health visit in the previous 3 years (n=3,460,932) (ii) all adult COVID-19 cases diagnosed before June 1, 2020 (n=15,203) and (iii) all hospitalized COVID-19 cases (n=2, 978).

### Potential bias from unmeasured confounding

To assess whether the association between HIV and COVID-19 mortality could be due to residual unmeasured confounding e.g. by socio-economic status, or unrecorded comorbidities, we calculated the E-value for an unmeasured confounder. The E-value for the analysis among all public sector patients was 3.70 (and 2.79 for the lower bound of the CI), suggesting that only a strong association between both HIV and low socio-economic status (or another confounder such as raised BMI), and low socio-economic status and COVID-19 death would account for all of the observed association between HIV and COVID-19 death. Most public sector patients have low socio-economic status with limited variability, and the effect of HIV on COVID-19 death was similar when restricting to the poorest subdistrict with the highest HIV prevalence in Cape Town.^15^ Similarly, quantitative bias analysis showed that the HIV-associated increased risk of COVID-19 death was unlikely due to confounding by raised BMI (Supplementary Appendix 4).

### Standardized mortality ratio

Among all laboratory diagnosed COVID-19 cases, there were 135 deaths among an estimated ∼520,000 PLWH in the province (260 deaths/million) and 786 deaths among 6.36 million people without HIV (124 deaths/million). The SMR for COVID-19 mortality in PLWH, relative to HIV-negative people was 2.39 (95% CI: 1.96; 2.86) and the attributable fraction of COVID-19 deaths due to HIV was 8.5% (95% CI: 6.1; 11.1).

## Discussion

In this analysis of nearly 3.5 million adults (16% PLWH) in South Africa we found an approximately two-fold increased risk of COVID-19 death in PLWH, irrespective of viral suppression, and a similar increased risk for patients with current tuberculosis. PLWH receiving TDF had a lower risk of COVID-19 death compared to those on other antiretrovirals, however these results may be confounded in the context of a public health approach with TDF in recommended first-line ART unless contra-indicated. While the HIV- and tuberculosis-associated increased risk of COVID-19 death may be over-estimated if there is residual confounding due to socio-economic status or unrecorded comorbidities, our results support considering people with HIV and tuberculosis as being at elevated risk of severe COVID-19. Nonetheless, despite a high burden of advanced HIV in the province, the attributable fraction of all deaths which could be ascribed to HIV was less than 10%.

While most case series of HIV and SARS-CoV-2 co-infection have not shown high risk of poor outcomes in PLWH, ^1-3,5-7^ cohorts of hospitalized PLWH with COVID-19 in London and New York have reported substantial morbidity and mortality including among patients with a suppressed viral load on ART.^18,19^ A comparison of hospitalized COVID-19 PLWH and people without HIV in New York suggested that COVID-19 outcomes may be worse in PLWH, but the study was under-powered to detect differences by HIV status.^20^ Absence of increased mortality risk in hospitalized patients with comorbidities may be explained by selection bias; risk factors for COVID-19 death may be attenuated by restricting to the subset of hospitalized patients who are already at high risk of mortality.^21^ It is therefore expected that for all comorbidities, the increased risk of death was progressively attenuated when restricting to cases (people with sufficiently severe symptoms to be tested) and hospitalized patients.

Similar to our findings, several studies have reported a high prevalence of comorbidities among PLWH with severe COVID-19.^3,6,7^ In our study, the overall high prevalence of diabetes in people with and without HIV, high proportion with poor diabetic control and elevated risks for COVID-19 compared to those reported from other countries are particularly concerning.^10^ The high prevalence of comorbidities in deceased PLWH suggests that the effect of HIV may at least partly be due to an increased risk of comorbidities at younger ages, possibly due to the “aging effect” of HIV.^2,7^ including those not recorded in WCPHDC such as cardiovascular disease. Persistent immune dysfunction may also play a role in severe COVID-19 despite viral suppression, and the hazard ratio point estimates for association with COVID-19 death were greater in immunosuppressed or viraemic PLWH, although the numbers of these patients with COVID-19 were small with wide CIs. Our finding that among COVID-19 cases in PLWH on ART those on TDF-based ART (vs. other therapies) were protected against COVID-death concurs with a recently published cohort of PLWH on ART from Spain.^13^ The protective effect of TDF in our setting may be over-estimated as TDF is only used in first-line therapy and contra-indicated in patients with poor renal function, however our analysis was adjusted for kidney disease and viral suppression. CD4 <200 cells/µl during admission was associated with COVID-19 death; this is likely due to the well-described lymphopenia in severe COVID-19 which is known to be prognostic of poor outcomes.^9^

We found both current and previous tuberculosis to be associated with COVID-19 death. While tuberculosis may exacerbate COVID-19 with impaired immune responses and increased angiotensin converting enzyme 2 receptor expression in respiratory epithelial cells, COVID-19 pneumonia may enhance tuberculosis progression.^22-24^ Nonetheless, since current tuberculosis itself causes death, it is difficult to disentangle the effects of COVID-19 vs tuberculosis disease on mortality *per se*.

To our knowledge this is the largest report on SARS-CoV-2 and HIV co-infected patients. Strengths include the study size using population-level data, laboratory confirmed SARS-CoV-2 diagnoses in all COVID-19 patients, inclusion of hospitalized and non-hospitalized cases and deaths, and coherence of associations from population (cohort study and SMR) to diagnosed to hospitalised patients. Being an observational study, limitations include potential under-ascertainment of comorbidities in routine administrative data, lack of data on other potential risk factors including socio-economic status and BMI, and relatively large numbers of PLWH without recent viral load or CD4 count results. In particular, patients with no recent information on disease control (e.g. HIV-RNA or HBA1c) may have less contact with health services and not reside permanently in the province, with under-ascertainment of outcomes.

## Conclusion

While our findings of a doubling of mortality risk in COVID-19 PLWH may over-estimate the effect of HIV on COVID-19 death due to the presence of residual confounding, PLWH should nonetheless be considered for inclusion as a high-risk group for COVID-19 management irrespective of virologic suppression, especially if they have other comorbidities.

## Data Availability

Data from the study is not available as was conducted on routine patient data and the requirement for informed consent was waived. Analysis code is available on request.

## Acknowledgements

We would like to acknowledge all patients in the Western Cape and to thank the Western Cape Department of Health Provincial Health Data Centre, the Western Cape Department of Health COVID-19 Outbreak Response Team, the Western Cape Communicable Disease Control sub-directorate and Western Cape health care workers involved in the COVID-19 response for their contributions to this report. We acknowledge funding for the Western Cape Provincial Health Data Centre from the Western Cape Department of Health, the US National Institutes for Health (R01 HD080465, U01 AI069924), the Bill and Melinda Gates Foundation (1164272), and the Wellcome Trust (203135/Z/16/Z).

## Notes

### Competing Interest Statement

The authors have declared no competing interest.

### Author Declarations

University of Cape Town Stellenbosch University

